# Hybrid surveillance and Bayesian modelling to estimate brucellosis incidence in a livestock-keeping population in northern Tanzania

**DOI:** 10.64898/2026.09.24.26363878

**Authors:** A.L. Holmes, W.A. de Glanville, V. Katiti, Â.J.F. Mendes, R.F. Bodenham, A. Lukambagire, B.T. Mmbaga, M.P. Rubach, J.A. Crump, S. Cleaveland, M. Viana, J.E.B. Halliday

## Abstract

Brucellosis is a globally distributed and widely under-appreciated zoonotic bacterial disease that causes infertility and abortion in livestock and febrile illness in humans, sometimes with debilitating sequelae. Brucellosis incidence estimates typically focus on acute presentations and range from 50 to 100 per 100,000 people per year for endemic settings, showing marked variation within and between countries. Estimating brucellosis incidence in low-resource settings is complicated by lack of access to and utilisation of health care and high-quality diagnostic testing for many people in rural communities. To estimate brucellosis incidence in a pastoralist population in northern Tanzania, we developed a Bayesian model that integrates multiple data sources including diagnostic test results from a hospital-based prevalence study performed in 2016-2017, test sensitivity and specificity, and data from a community-based healthcare utilisation survey in 2019-2020. We show that this population experienced a high incidence of brucellosis with wide uncertainty, with estimates of 387.6 (95% HPDI: 103.3 - 903.5) illnesses per 100,000 people per year for the total population, 199.7 (95% HPDI: 16.2 - 584.6) for individuals aged >15 years, and 632.4 (95% HPDI: 180.1 - 1379.4) for children aged ≤15 years. These incidence estimates were among the highest in the world to date. The wide density intervals for these incidence estimates reflect uncertainty in the underlying parameter values used. Sensitivity analyses show that healthcare utilisation influenced incidence estimates the most. This result indicates that accounting for low levels of healthcare utilisation is essential to avoid under-estimating disease incidence. Our findings illustrate that brucellosis is a frequent cause of illness in this population. Increased access to treatment, prevention, and accurate diagnostic testing would greatly improve understanding of brucellosis epidemiology and the health of many people living in similar at-risk communities.

## Introduction

Brucellosis is a neglected zoonotic disease caused by infection with bacteria belonging to one of several *Brucella* species. Globally, most human brucellosis infections occur in low- and middle-income countries and are linked to direct animal contact or the consumption of *Brucella*-contaminated animal-source foods (1). Acute human brucellosis generally presents as a non-specific febrile illness, requiring laboratory tests for accurate diagnosis. Febrile illness is observed in approximately 78% of presentations with acute brucellosis, but other symptoms and signs are highly variable (2). Accurate diagnostic tests are often unavailable in endemic countries, and widely used tests often have poor sensitivity and specificity (3–7). In the absence of accurate diagnostic tests in low-resource settings, human brucellosis may commonly be misdiagnosed as other, apparently more common or better-known causes of fever, such as malaria (8, 9). The use of diagnostic tests with poor specificity can also result in over-diagnosis of brucellosis (5). The resulting uncertainty about the incidence of human brucellosis in many low- and middle-income countries has important implications for disease prioritisation and resource allocation (7).

Human brucellosis is preventable through vaccination and test-and-slaughter of livestock, which have contributed to *Brucella spp*. elimination in some high-income countries (10–12). However, such measures are logistically challenging and expensive to implement (13). To assess whether candidate interventions are cost-effective in currently endemic settings, high-quality estimates of human brucellosis incidence are needed to estimate the potential impact of interventions (14), particularly in countries that currently apply minimal or no brucellosis control in livestock (15).

Cohort studies in which individuals are followed over time and new disease events are detected using accurate diagnostic tests provide the gold standard methodological approach for incidence estimation. However, such studies require considerable resources to implement, particularly in low-resource settings and may be impractical beyond specific studies in high-risk populations (16). An alternative, lower-cost approach is to undertake ‘hybrid surveillance’, linking health facility-based brucellosis detection with healthcare utilisation surveys. This allows the estimated incidence of disease derived from health facility-based data to be adjusted by the proportion of people in the health facility’s catchment area that are expected to seek care at the sentinel facility if they develop illness, while also adjusting for diagnostic test performance and enrolment levels (17). Similar approaches have been used to estimate the population-level incidence for a range of infectious diseases in low- and middle-income settings (17–21), including human brucellosis (18).

Brucellosis is endemic in Tanzania (22), but few estimates of human disease prevalence or incidence exist. Within the Ngorongoro Conservation Area (NCA), an area in northern Tanzania in which pastoralism is practiced, the prevalence of probable or confirmed brucellosis among febrile persons presenting for care at the largest health facility was estimated as 6% in 2016-2017 (23). A previous study that estimated human brucellosis incidence with a hybrid surveillance approach reported 33 illnesses per 100,000 people per year for the more urban population in and around the town of Moshi in northern Tanzania (18).

Northern Tanzania has a diversity of livestock production systems (24), and since animal *Brucella* infection risk is often strongly linked to the type of livestock production system, livestock infection prevalence likely varies considerably between systems (25–27). The peri-urban areas around Moshi for which previous human brucellosis incidence estimates were derived are characterised as smallholder livestock production where people keep small numbers of cattle and small ruminants, often in intensive zero-grazing systems (24). The prevalence of livestock brucellosis in such systems is often low (28), and typically much lower than in systems characterised by more extensive livestock production, such as pastoralist systems (12, 27, 29). Human brucellosis risk, particularly in rural areas, is linked to brucellosis prevalence in the local livestock population (30, 31) and to cultural practices that influence the risk of exposure to contaminated animal-source foods such as raw dairy products (32). Therefore, the incidence of human brucellosis is likely heterogeneous in endemic countries (15, 33, 34), with risks of infection particularly high in pastoralist settings (16, 32–35).

Here, we use a hybrid surveillance approach and a Bayesian latent process model that integrates diagnostic results from serological tests and blood culture while accounting for their sensitivity and specificity, with estimates of health care utilisation to estimate the population-level incidence of brucellosis for adults, children, and the combined general population living within the NCA. We further examine the sensitivity of incidence estimates to input parameters and evaluate how the complexity of the model and methods of estimating healthcare utilisation affect the incidence estimate.

## Methods

### Hospital-based fever study

#### Study health facility and population

The NCA is a large mixed land use area of 8,100 km^2^ in which pastoral livelihoods are practiced alongside wildlife conservation and tourism (36). The local, predominantly Maasai, population keep cattle, sheep, and goats that are managed extensively. Consistent with the epidemiology in most of mainland Tanzania, *Brucella* was expected to be circulating endemically in livestock within the NCA (37), and was essentially uncontrolled. The total human population of the NCA was 93,136 people as of 2012, the most recent census conducted prior to the foundational prevalence study (38).

At the time of the prevalence study, Endulen Hospital was the only hospital within the NCA. There were also smaller health centres and dispensaries within and outside the NCA at which people with febrile illness could seek care. Although people from outside the NCA could and did seek care at Endulen Hospital, these numbers are typically small (23).

#### Human brucellosis case definition

Human brucellosis case data were generated for patients enrolled at Endulen Hospital from 15 August 2016 through 11 October 2017, with sampling taking place on a total of 259 working days (23). Individuals seeking care at the outpatient department of Endulen Hospital aged two years or older with reported fever within the past 72 hours or with a tympanic temperature of ≥38.0 °C at presentation were enrolled.

At acute phase sampling, blood was collected from participants and inoculated into two 10mL BacT/ALERT (BioMerieux, Marcy-l’Étoile, France) aerobic blood culture bottles for continuously monitored incubation and one 10mL Castaneda media (prepared at the Animal and Plant Health Agency (APHA), Weybridge, UK) bottle for incubation and manual monitoring as described elsewhere (23). Standard methods were used for identifying bacterial isolates (39, 40). Presumptive *Brucella* spp. were isolates of Gram-negative coccobacilli that had positive reactions for urease, catalase, and oxidase. Identification was confirmed and species typed using multiplex conventional PCR and multilocus sequence typing at the APHA, UK (41). Participants were also visited in their homes four to six weeks after enrolment for convalescent-phase sampling. At both acute- and convalescent-phase sampling events, up to 10mL of blood was collected into a plain vacutainer from which serum was separated. Serum was tested for the presence of antibodies to *Brucella* spp. using a serum agglutination test (SAT) performed at the APHA, UK (41). Full details of the foundational brucellosis prevalence study are reported elsewhere (23).

A confirmed case of acute human brucellosis was defined by isolation from blood culture of *Brucella* spp. or a four-fold or greater rise in *Brucella* SAT antibody titre between acute and convalescent samples, in a febrile person. Probable cases of brucellosis were defined by a SAT antibody titre >160 in the acute or convalescent phase serum of a febrile person (23). For this study both confirmed and probable brucellosis cases contribute to the estimate of brucellosis incidence; results from the acute and convalescent phase sample were combined and an individual classified as ‘positive’ by serology if they had an SAT antibody titre >160 in their acute sample, convalescent sample or both.

#### Healthcare utilisation survey

A household healthcare utilisation survey (HCUS) was conducted to estimate the proportion of people residing in the NCA who would attend Endulen Hospital if they developed febrile illness (later included in the incidence estimate model as the parameter “utilisation”, or *U*). The HCUS was conducted within the NCA from 7 November 2019 through 17 March 2020 after piloting in October and early November 2019. The target sample size was 679 households. Details of the boma and household definitions applied for the study are given in the Supplementary Materials. This sample size was calculated to allow estimation of a conservative household-level prevalence of 50% for a range of outcomes with 10% precision at the 95% confidence level and assuming a conservative intra-cluster correlation at the sub-village level of 40% (42), and 15 households surveyed per cluster. Sample size estimation was conducted using standard formulae (43).

Within selected sub-villages, a list of all bomas present was generated with a group of elders. A boma was a unit of one or more related families that live together within a single compound. Within each sub-village, 15 bomas were randomly selected and within each boma, a single household was randomly selected. If the boma or household respondent refused to participate or did not respond, a back-up boma was randomly selected and the process repeated. All random selection (of sub-villages, bomas, and households) was performed using a random number generator.

Within selected households, all available adult members (>18 years for the purposes of consent), excluding those who were the children of the household head, were invited to participate in the HCUS. These adults were typically the male household head and his wife or wives, who were assumed to be the main decision makers for the household. Sections of the survey related to household-level decision making, or that required knowledge about specific individuals in the household, were administered with all available eligible adults in the household. If one or more household decision maker was temporarily unavailable, a return visit within 24 hours to conduct the survey was arranged. When decision makers were unavailable for more than 24 hours (e.g., because of travel), the survey was conducted with all decision makers currently present.

Household location coordinates were collected using a handheld GPS device (Garmin, eTrex 10). Questionnaire data were collected using Open Data Kit (ODK) on handheld tablets. The data domains covered by the survey tool and analysed for this study included household member demographics, decision making and actions taken around both hypothetical febrile illness among household members, reported febrile illness among household members in the 14 days before survey administration, and associated actions. Further details of the questions asked to generate data used for this study are given in the Supplementary Materials (Table S1-S3).

#### Alternative approaches to quantify healthcare utilisation

The model parameter *U* represented the proportion of people in the study area who would attend Endulen Hospital if they developed a febrile illness consistent with brucellosis (see additional details below). Given the non-specific clinical manifestations of brucellosis, the descriptions used here focus largely on febrile illness and were intended to describe and capture acute presentations with brucellosis. Previous applications of the hybrid surveillance approach for brucellosis estimated this parameter using responses to the question “To which health facility would you go if you were unwell with a fever lasting three or more days?” (18). The framing of this question provided at least two important sources of uncertainty. The first was the extent to which reporting of hypothetical healthcare seeking for fever reflects actual health seeking for fever. The second was whether health seeking behaviour for fever can be considered the same as healthcare seeking behaviour for brucellosis. To address this, we quantified *U* with three different approaches. The first approach (*U1*) was used for the final (full) model, as it was the most comparable to other similar studies conducted previously in northern Tanzania (18, 21, 44), and the other two approaches were used to assess how survey methodology affects the utilisation and consequently incidence estimates obtained. All survey questions were asked of four age groups: children aged <2 years, children ≥2 <5 years, children ≥5 and ≤15, and anyone aged 16 years or older. For modelling, data from the three youngest age groups were aggregated to obtain parameter estimates for individuals ≤15 years and under as a single group.

#### Method 1: Hypothetical care seeking for febrile illness (*U1*)

First, household respondents were asked “what would you do if someone in this household developed a febrile illness lasting more than three days?” Respondents were asked to provide their three most likely courses of action in each scenario. If attending a health facility was among these choices, respondents were asked which named health facility that they would attend. The proportion of respondents that named Endulen Hospital as a named health facility that they would attend, if visiting a health facility at all was listed as one of the top three actions taken, in response to febrile illness of more than three days duration in a household member was used as an estimate of healthcare seeking behaviour.

#### Method 2: Reported healthcare utilisation for febrile illness (*U2*)

Second, respondents were asked “Has any person in this household had a febrile illness in the past 14 days?” If the response was yes, respondents were asked if they sought care at a health facility for this illness and which health facility, or facilities, they used. The proportion of febrile people that reported seeking healthcare at Endulen Hospital in the 14 days prior to survey administration was used as an estimate of actual healthcare seeking behaviour.

#### Method 3: Detailed brucellosis-like scenario (*U3*)

Scenarios in which a hypothetical female child continued to feel unwell after 10 days and 3 months were explained and respondents were asked what they would do at each of these three time points: initial onset of febrile illness, febrile illness of 10-day duration, and febrile illness of 3-month duration. If respondents reported that they would seek healthcare at one or more of these time points, they were asked which health facility they would attend. The total proportion of respondents who reported that they would have sought care at Endulen Hospital at any time point was calculated and the responses to these questions were used to estimate the proportion of people who would attend Endulen Hospital. This scenario was intended to describe the acute or sub-acute febrile illness marked by intermittent or remittent fever which can persist for weeks or months with brucellosis (11). Estimates of *U3* were only applied to estimate brucellosis incidence in children.

Full details of the survey questions asked to obtain healthcare utilisation estimates are given in the supplementary materials (Supplementary Table 1-3). To account for clustering in survey delivery, raw proportions and 95% CI obtained from the survey responses were adjusted to account for clustering at the sub-village level using the *survey* package v. 4.4-2 (45) in R version 4.4.2 (46). These adjusted point estimates and confidence intervals were used to define the values and prior distributions used for modelling.

**Table 1:**
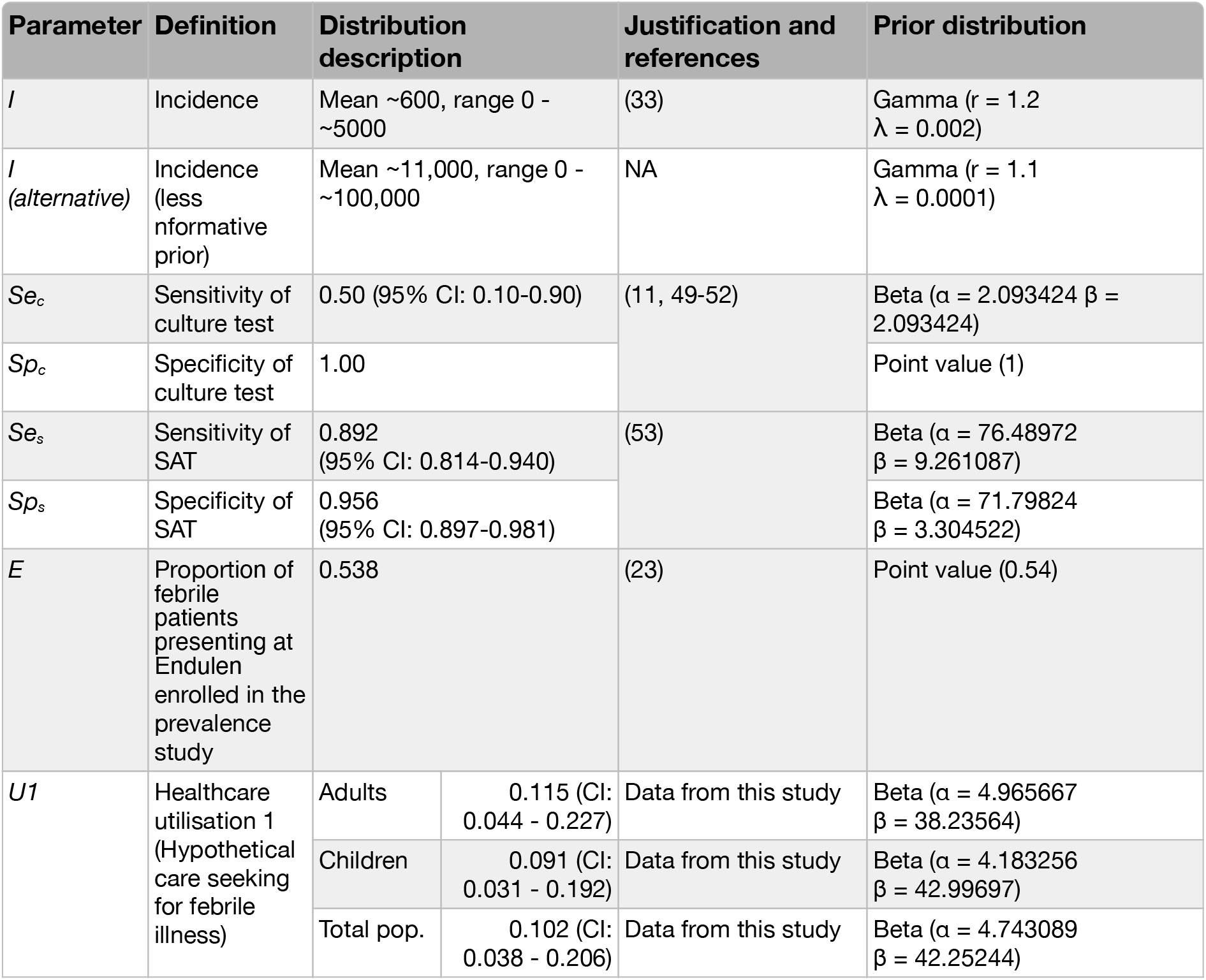
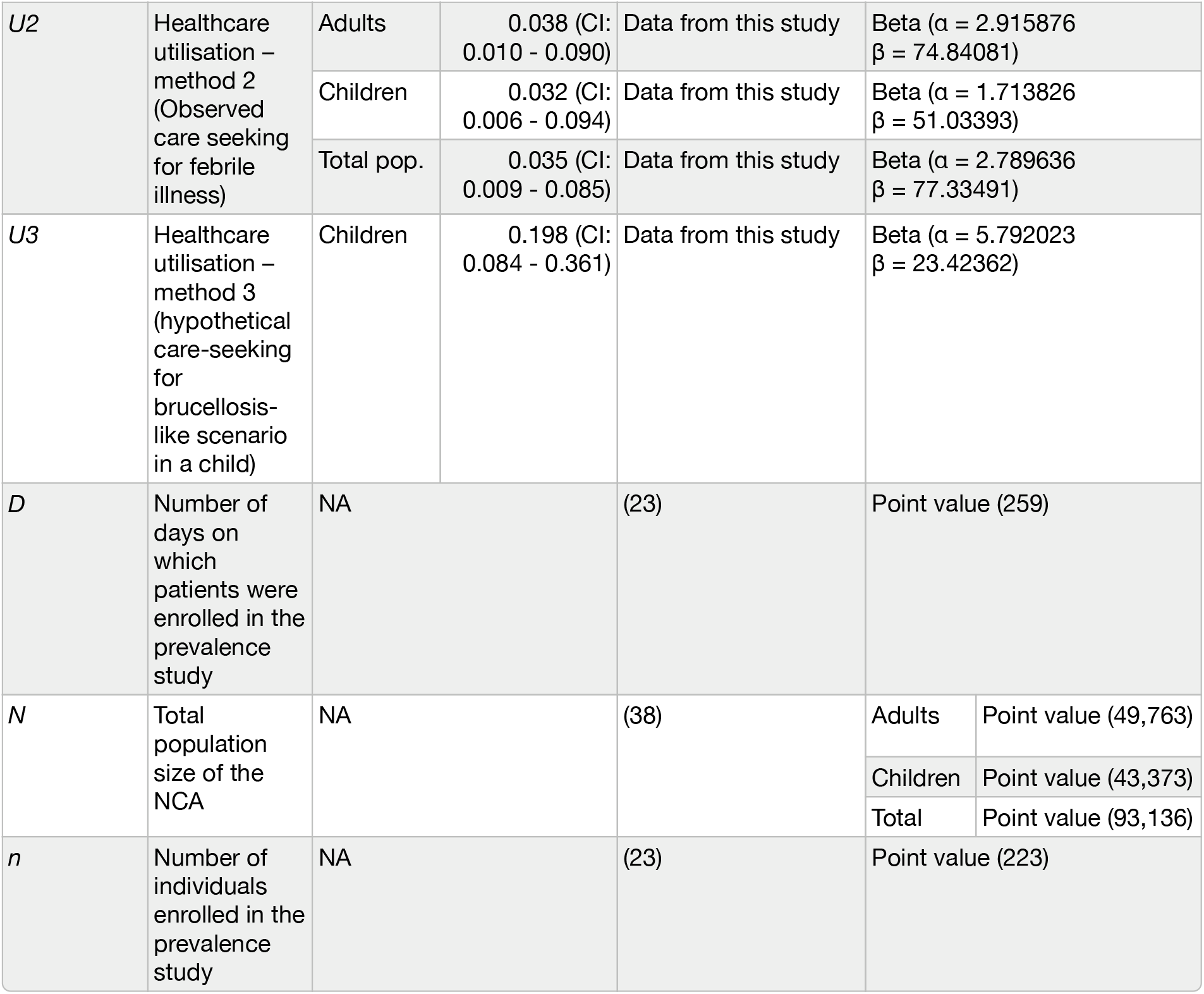
Summary of data values and prior distributions used to define parameters for estimation of brucellosis incidence in the population of the Ngorongoro Conservation Area, Tanzania, 2016-2017.

| Parameter | Definition | Distribution description |  | Justification and references | Prior distribution |
| --- | --- | --- | --- | --- | --- |
| $I$ | Incidence | Mean ~600, range 0 - ~5000 | | (33) | Gamma ($r = 1.2$<br>$\lambda = 0.002$) |
| $I$<br>(*alternative*) | Incidence (less nformative prior) | Mean ~11,000, range 0 - ~100,000 | | NA | Gamma ($r = 1.1$<br>$\lambda = 0.0001$) |
| $Se_c$ | Sensitivity of culture test | 0.50 (95% CI: 0.10-0.90) | | (11, 49-52) | Beta ($\alpha = 2.093424$ $\beta = 2.093424$) |
| $Sp_c$ | Specificity of culture test | 1.00 | | | Point value (1) |
| $Se_s$ | Sensitivity of SAT | 0.892 (95% CI: 0.814-0.940) | | (53) | Beta ($\alpha = 76.48972$<br>$\beta = 9.261087$) |
| $Sp_s$ | Specificity of SAT | 0.956 (95% CI: 0.897-0.981) | | | Beta ($\alpha = 71.79824$<br>$\beta = 3.304522$) |
| $E$ | Proportion of febrile patients presenting at Endulen enrolled in the prevalence study | 0.538 | | (23) | Point value (0.54) |
| $U1$ | Healthcare utilisation 1 (Hypothetical care seeking for febrile illness) | Adults | 0.115 (CI: 0.044 - 0.227) | Data from this study | Beta ($\alpha = 4.965667$<br>$\beta = 38.23564$) |
| | | Children | 0.091 (CI: 0.031 - 0.192) | Data from this study | Beta ($\alpha = 4.183256$<br>$\beta = 42.99697$) |
| | | Total pop. | 0.102 (CI: 0.038 - 0.206) | Data from this study | Beta ($\alpha = 4.743089$<br>$\beta = 42.25244$) |

|  |  |  |  |  |  |  |
| --- | --- | --- | --- | --- | --- | --- |
| *U2* | Healthcare utilisation – method 2 (Observed care seeking for febrile illness) | Adults | 0.038 (CI: 0.010 - 0.090) | Data from this study | Beta ($\alpha$ = 2.915876<br>$\beta$ = 74.84081) | |
| | | Children | 0.032 (CI: 0.006 - 0.094) | Data from this study | Beta ($\alpha$ = 1.713826<br>$\beta$ = 51.03393) | |
| | | Total pop. | 0.035 (CI: 0.009 - 0.085) | Data from this study | Beta ($\alpha$ = 2.789636<br>$\beta$ = 77.33491) | |
| *U3* | Healthcare utilisation – method 3 (hypothetical care-seeking for brucellosis-like scenario in a child) | Children | 0.198 (CI: 0.084 - 0.361) | Data from this study | Beta ($\alpha$ = 5.792023<br>$\beta$ = 23.42362) | |
| *D* | Number of days on which patients were enrolled in the prevalence study | NA |  | (23) | Point value (259) |  |
| *N* | Total population size of the NCA | NA |  | (38) | Adults | Point value (49,763) |
|  |  |  |  |  | Children | Point value (43,373) |
|  |  |  |  |  | Total | Point value (93,136) |
| *n* | Number of individuals enrolled in the prevalence study | NA |  | (23) | Point value (223) |  |

### Incidence estimates

To estimate the incidence of brucellosis in the NCA, we fitted a Bayesian latent process model to integrate the different data streams and estimate uncertainty, expanding on an approach used previously (47). The model was fitted to the data for adults, children, and the total population (i.e. all ages combined). In this model, the probability that a febrile person *i* (in this context, someone presenting to Endulen with febrile illness) of age group *j* had brucellosis (*p*_*i,j*_) was defined as:

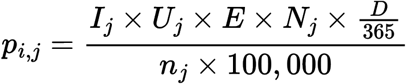

where *I*_*j*_ was the incidence (number of incident cases per 100,000 people per year) of brucellosis in age group *j*. The prior for *I*_*j*_ was defined based on the belief that incidence is often underestimated. As such, it was described by a gamma distribution that produced mean values of approximately 600, slightly higher than the highest estimates found in the literature to date, e.g. (33), but also acknowledging uncertainty by permitting incidence values ranging from 0 up to approximately 5000 (Table 1). To ensure the estimates were not being constrained by the prior, the model was also run with an alternative and less informative prior (mean = 11,000, range: 0 - 100,000; Gamma, r = 1.1, λ = 0.0001).

In all models, point values used for the parameters *E* (the proportion of people enrolled in the prevalence study) and *D* (the number of days the study ran for) were taken from the prevalence study and the point values for *N* (the total population size) were taken from census data (38).

### Likelihood of diagnostic test data

The process by which diagnostic test data, that is, the results of the SAT serology (*S*_*i,j*_) and culture (*C*_*i,j*_) testing for each febrile person *i* in age group *j*, informed the estimates of incidence were defined by Bernoulli distributions with probabilities *p*_*s*_ or *p*_*c*_ of being positive for each test, respectively:

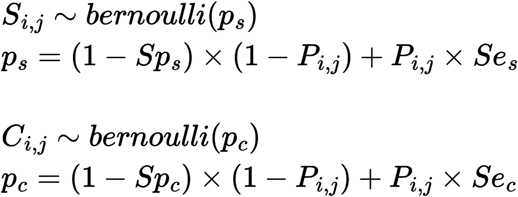

*Sp*_*s*_ and *Sp*_*c*_ were the specificity of SAT serology and culture, respectively, and *Se*_*s*_ and *Se*_*c*_ were the sensitivity of these tests. Beta distributions for the sensitivity and specificity parameters were used as priors. These were defined by transforming literature informed mean values into beta distribution parameters using the *BetaExpert* function from the package *prevalence* (48) (Table 1). A specificity of 100% was assumed for culture, and a prior distribution allowing a wide range of values was specified for sensitivity was specified, reflecting overall lack of evidence on the sensitivity of this test in this or similar populations (Table 1). *P*_*i,j*_ was the estimated brucellosis case status (case or non-case) for each individual *i* in age group *j*, which was drawn from a Bernoulli distribution defined by *p*_*i,j*_:

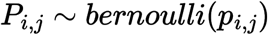

The modelled case status of each participant in the prevalence study was estimated based on their culture and SAT results and the sensitivity and specificity of these tests. Most participants had both culture and SAT result data. For participants with complete SAT data but no culture results, these missing data were predicted within the model as part of the joint modelling process. For the full model run, parameters *I, U, Se*_*c*_, *Sp*_*c*_, *Se*_*s*_ and *Sp*_*s*_ were specified by prior distributions to account for their uncertainty. Details of the specification of all parameters and simplified model versions considered are given in Table 1 and below.

The Bayesian models were run in JAGS using the package ‘*rjags’* version 4-17 (54) for 100,000 iterations on three separate chains with 50,000 iterations for burn in. Convergence was checked by visual inspection of the trace plots and Rhat values between chains, and model goodness of fit was verified with posterior checks. Additionally, predictions of brucellosis case status for individuals with each observed combination of diagnostic test results were checked to confirm that the model predicted infection status at proportions consistent with our expectation of each test’s sensitivity and specificity (predicted probabilities of being a brucellosis case for each combination of test results can be found in Supplementary Table 5).

#### Model simplification and sensitivity analysis

To understand the relative influence of the model parameters included and their specification (i.e., comparing specification as point value versus a prior distribution to introduce uncertainty) upon the incidence estimates, a series of simplifications of the full model (FM) were run, following the example of Cutting et al. (2022). For model simplification 1 (MS1), instead of a prior distribution, U1 was provided as a point estimate with the value equivalent to the mean for *U1* for each population (Table 1), essentially assuming no uncertainty with *U1*. MS2 included the prior for *U1* but removed diagnostic test sensitivity and specificity from the model, effectively assuming 100% sensitivity and specificity for both tests. For MS3, the utilisation parameter *U* was removed entirely, effectively assuming 100% healthcare utilisation, but priors for test sensitivity and specificity were retained. For MS4, both test sensitivity and specificity and *U* were removed, assuming perfect test performance and 100% healthcare utilisation. For models not accounting for test sensitivity and specificity, the diagnostic data informing the model were the brucellosis case status data only (confirmed and probable cases as compared to individuals who did not have brucellosis) rather than the separate results of the SAT and culture tests.

Finally, to directly compare estimates from this setting with other published values from different regions, two deterministic calculations were performed. First, an incidence calculation highly similar to that employed by Carugati et al (2018) was performed (MS5), where incidence was calculated as: I = (Cases ^*^ (1/N) ^*^ (365/D) ^*^ (1/E) ^*^ (1/SAT_Sensitivity) ^*^ (1/U1)) ^*^ 100000. For this calculation, the *U1* value for healthcare utilisation was used and only the SAT data from the Endulen population were used (not also the culture data). This calculation does not include the paired “sera multiplier”, the “referral adjustment multiplier”, or the “blood drawn multiplier that were included in the Carugati estimate, as these parameters were not captured in the design of this study. Finally, a ‘crude’ incidence estimate (MS6) was calculated as (I = (Cases / (N ^*^ E ^*^ D / 365)) ^*^ 100,000).

The parameter *U* corresponds to the probability of healthcare seeking when febrile for a given age group. For all the models described above, the values and priors for *U1* specified in Table 1 were used. To determine the influence of the alternative values obtained for the utilisation parameter on the estimate of incidence, variations of the full model were run with each of the three alternative values: *U1, U2* and *U3. U2* and *U3* were given beta prior distributions defined by the survey-design-adjusted point estimates and 95% confidence intervals using the *betaExpert* function from the R package *prevalence* v. 0.4.1 (48) (Table 1), as for *U1*.

#### Research clearance and ethics

Approval to conduct the foundational prevalence study and healthcare utilisation study was granted by the Tanzania Commission for Science and Technology, Tanzania Wildlife Research Institute and the Ngorongoro Conservation Area Authority. Ethical approval was granted by the Kilimanjaro Christian Medical Centre (KCMC) University Research Ethics Committee (698 and 829); National Institute of Medical Research (NIMR), Tanzania (NIMR/HQ/R.8c/Vol. I/1140 and NIMR/HQ/R.8a/ Vol. IX/2079) University of Otago Human Ethics Committee (H17/052), and University of Glasgow College of Medical, Veterinary and Life Sciences Ethics Committee (200140149). The research was performed in accordance with the guidelines and regulations prescribed by the above organisations. Written informed consent for study participation was obtained from each participant and/or their legal guardian, using forms translated into Swahili and verbal translation into Maa when needed.

## Results

### Healthcare utilisation survey

Surveys gathered data on healthcare utilisation from 343 households in 20 sub-villages in and immediately adjacent to the NCA. Sampled households comprised 2,056 people. The target sample size was not achieved due to restrictions on field work imposed in response to the COVID-19 pandemic in 2020. The spatial distribution of sampled households in the NCA in relation to Endulen Hospital and the human population density within the NCA is shown in Figure 1.

**Figure 1:**
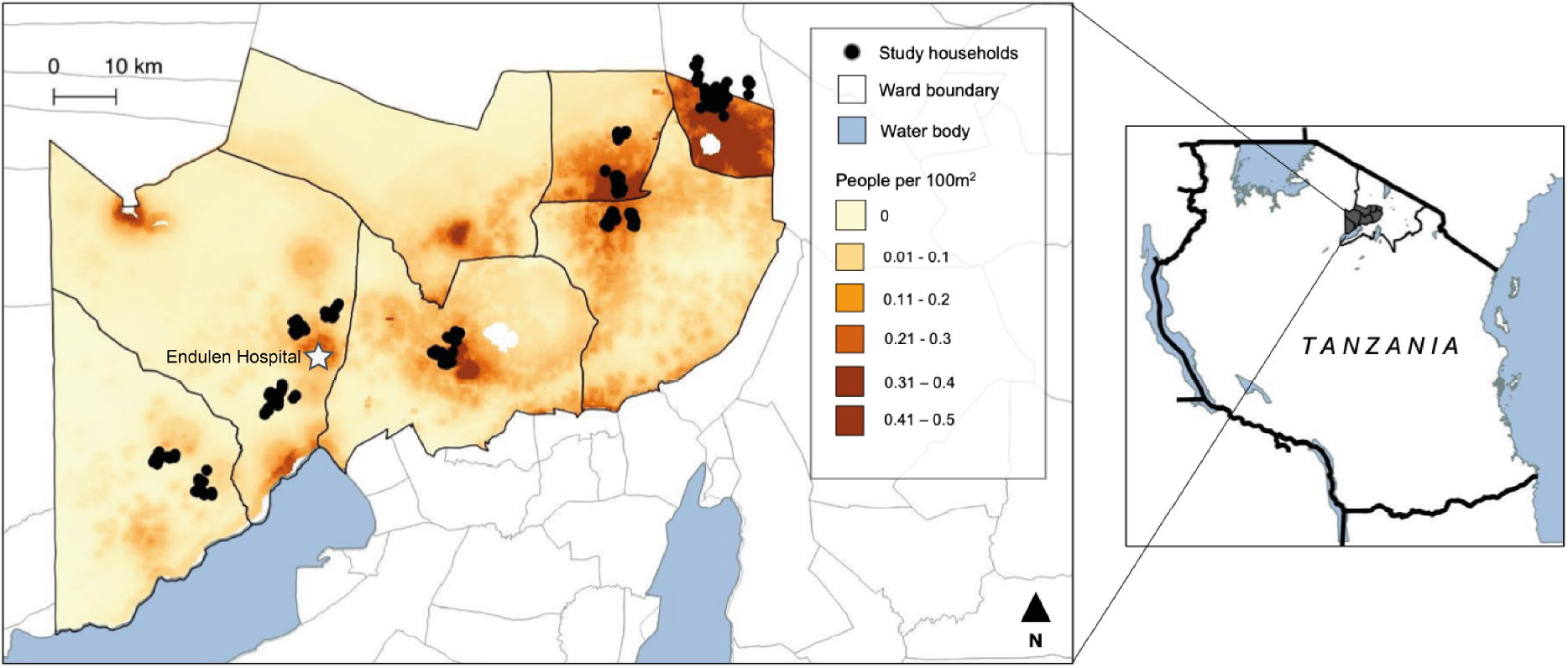
Location of households sampled in the healthcare utilisation survey performed within the Ngorongoro Conservation Area, northern Tanzania, 2019-202. Sampled household locations are indicated by black dots. The human population density of the study area (wards within the Ngorongoro Conservation Area) is indicated by the map shading (source data from worldpop.org (55)). Outlined polygons indicate ward administrative boundaries (source data from gadm.org (v. 4.1)). The location of Endulen hospital is marked with a white star and text label. Map generated in QGIS v. 3.22 (56).

**Figure 2:**
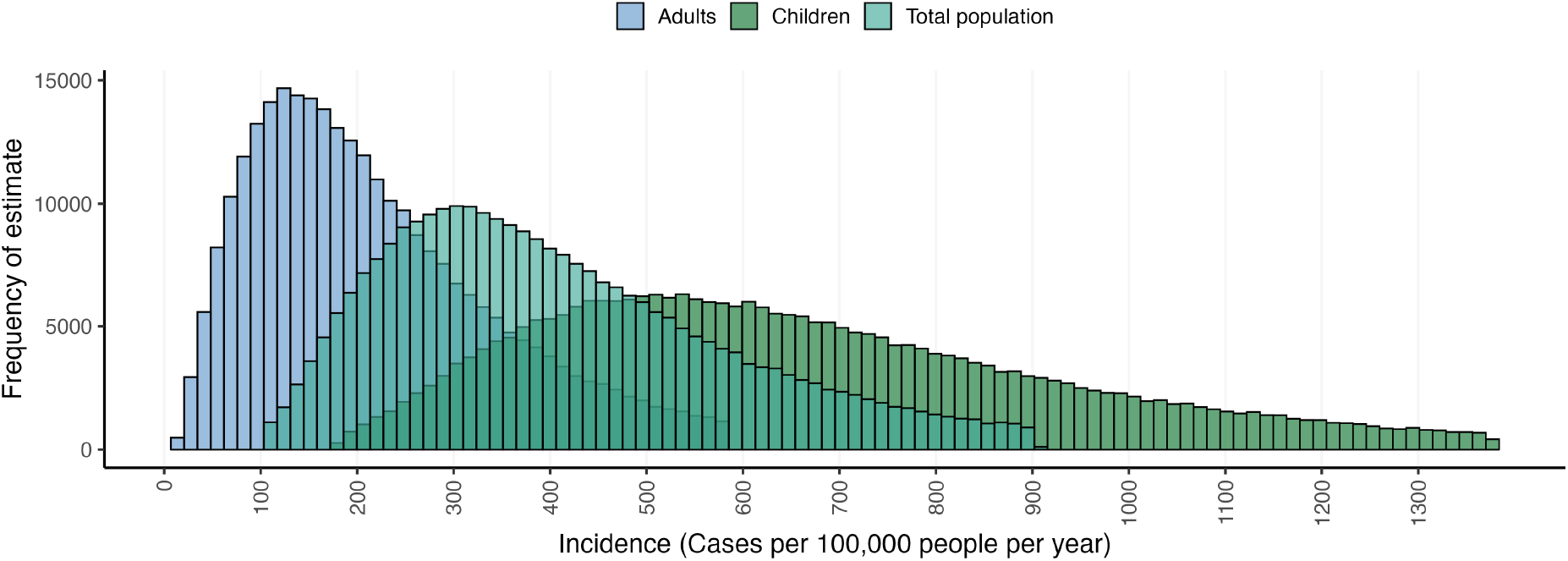
Posterior distributions of human brucellosis incidence estimates for the Ngorongoro Conservation area, 2016-2017. Only the 95% highest posterior density interval for each population is shown. The distributions for specific populations are indicated in colour as adults (blue), children (green), and the combined adult and child population (teal).

### Healthcare utilisation estimates

The estimates of healthcare utilisation at Endulen hospital varied between age groups and measurement methods (Table 1). When asked about hypothetical care seeking if a family member developed a fever lasting more than three days, the overall estimate of the proportion of individuals who would attend Endulen Hospital as one of their top three choices of healthcare facility was 0.102 (*U1*) (Table 1). In comparison, the equivalent estimate obtained from questions about reported healthcare seeking behaviour in response to actual recent febrile illness (*U2*) were substantially smaller, with an estimate for the total population of 0.035 (Table 1). For the specific scenario of a 4-year-old girl with persistent febrile symptoms and brucellosis-like illness the proportion of children who would present to Endulen was estimated as 0.198 (*U3*) (Table 1).

### Incidence model

The raw prevalence was 14 cases out of 229 tested individuals, with 3 cases in adults and 11 in children (23). The median incidence of brucellosis in the total population estimated using the full model and *U1* healthcare utilisation estimates was 387.6 cases per 100,000 people per year (95% highest posterior density interval (HPDI): 103.3 - 903.5). The case distribution was not even across the population (Figure 2); brucellosis cases were almost three times more likely in children, with a median incidence of 632.4 (95% HPDI: 180.1 - 1379.4) per 100,000 people per year, compared to 199.7 (95% HPDI: 16.2 - 584.6) per 100,000 people per year in adults (Figure 2). Additional plots of the estimates for each age group independently are available in Supplementary Figure 2. The model run with the alternative (less informative) prior for *I* showed a larger skew, leading to a higher median incidence estimate due to a small number of very high estimates that were considered unlikely, but the peaks remained comparable to the model with a more informative prior. The point estimates and HPDI values for all models are available in Supplementary Table 4, and the un-truncated histograms for the full model with both prior specifications can be found in Supplementary Figures 1 and 3.

### Sensitivity of incidence estimates to test performance and healthcare utilisation

The median incidence estimates obtained for the total population were similar between the full model and model simplifications (MS) 1 and 2, but the range of the HPDIs varied substantially, with the model using a point estimate for *U* (MS1) showing a much smaller range compared to those including a prior to capture uncertainty in *U* (Full model and MS2) (Figure 3). MS3 and MS4 generated much lower incidence estimates than the models including healthcare utilisation. The point estimate obtained using almost all parameters included in the full model in a deterministic (i.e., non-Bayesian) process (MS5) was similar to but lower than the median values obtained using the full model and first two simplifications. The simplest deterministic incidence calculation (MS6), which included neither *U* nor test sensitivity and specificity, provided a much lower estimate very similar to the median values obtained with the Bayesian models that did not include any terms for healthcare utilisation (MS3 and MS4) (Figure 3). The distributions in Figure 3 are truncated to the 95% HPDI for legibility, with an un-truncated version available in the supplementary materials (Figure S4). All model estimates and HPDI values can be found in Table S4, and comparisons of model simplifications for child and adult populations specifically are shown in Figures S5-6.

**Figure 3:**
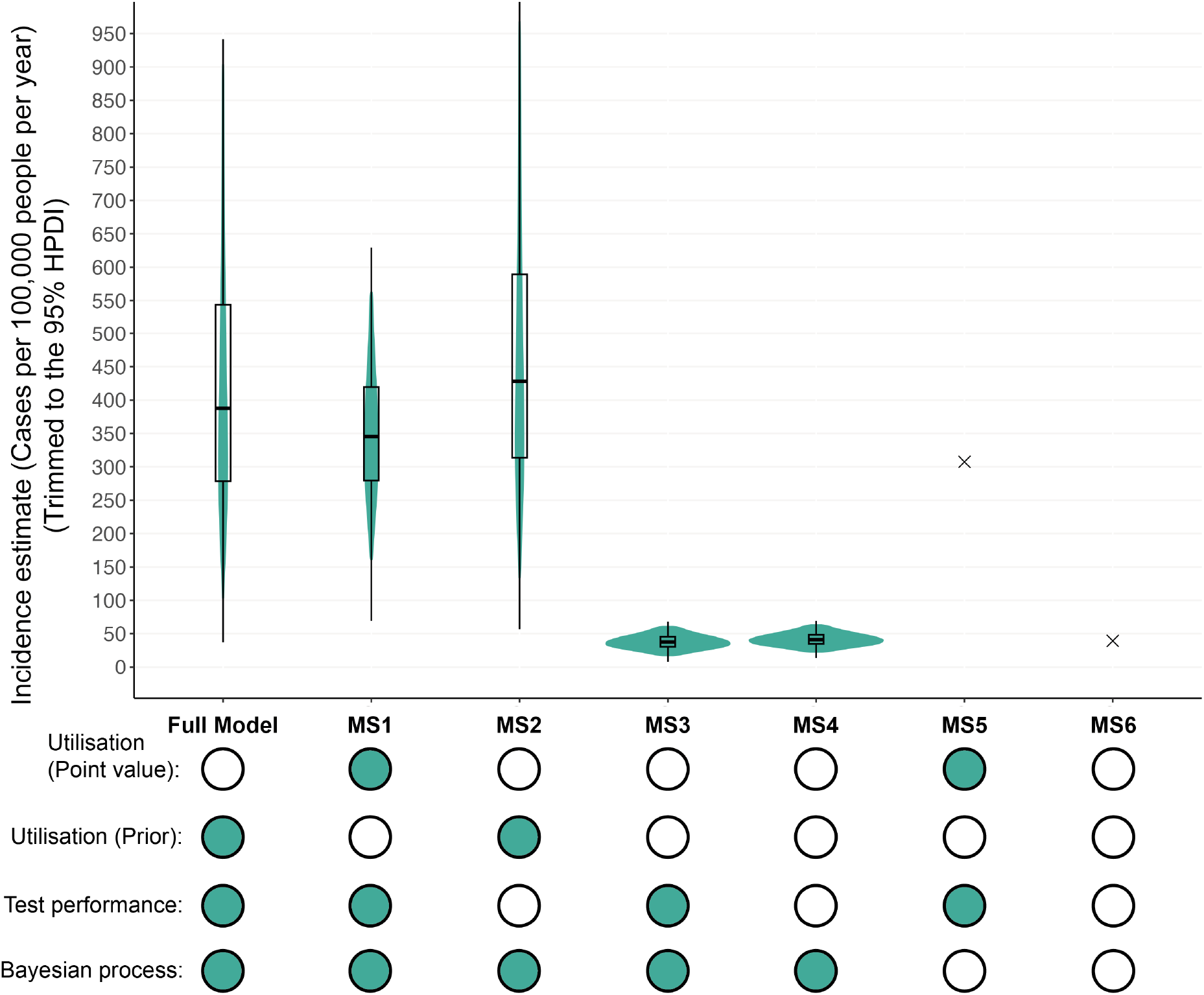
Posterior distributions of human brucellosis incidence estimates for the Ngorongoro Conservation area, 2016-2017, comparing estimates from the full model and simplified models. The 95% HPDIs for each model are presented in violins, with the boxplots indicating the median value and upper and lower quartiles. The estimates obtained from deterministic models (MS5 and MS6) are indicated by crosses on the graph. Circles below each model indicate whether that parameter is included in the model, with filled circles indicating that the parameter is included and empty circles indicating that the parameter is not included. Additionally, the lower line of circles differentiates estimates from Bayesian (filled circles) and non-Bayesian (empty circles) calculations. The full model includes healthcare utilisation and the test performance parameters (sensitivity and specificity) as prior distributions. Model simplification 1 (MS1) uses a single point value rather than a distribution for healthcare utilisation. Model simplification 2 (MS2) uses a prior distribution for utilisation but removes test performance parameters from the model. Model simplification 3 (MS3) removes healthcare utilisation entirely but retains test performance parameters. Model simplification 4 (MS4) removes test performance and utilisation parameters but remains a Bayesian process. MS5 includes a point value for utilisation and a correction for SAT sensitivity but is a deterministic rather than Bayesian process. MS6 is the simplest model, not accounting for either utilisation or test performance. All models summarised here were run using the estimate for U1 and its associated uncertainty as appropriate.

### Influence of alternative estimates of healthcare utilisation on incidence

Running the full model with different estimates of the healthcare utilisation parameter changed the final estimates and levels of uncertainty, but the HPDI of all estimates overlapped with each other within populations (adults, children and the total population) (Figure 4). Models using *U2*, which captured actual rather than hypothetical healthcare seeking behaviour (and had lower values for the utilisation parameter as compared to *U1* and *U3*, Table 1), produced the highest incidence estimates. In contrast, *U3*, which captured a detailed hypothetical scenario about a four-year-old girl (and was only applied to estimate incidence in children) provided an estimate that was slightly lower than that obtained using *U1* (Figure 4). Median incidence estimates for the population of children were 632.4 cases per 100,000 people per year using *U1*, 1035.1 using *U2* and 412.8 using *U3*. All model estimates and HPDI values can be found in Table S4.

**Figure 4:**
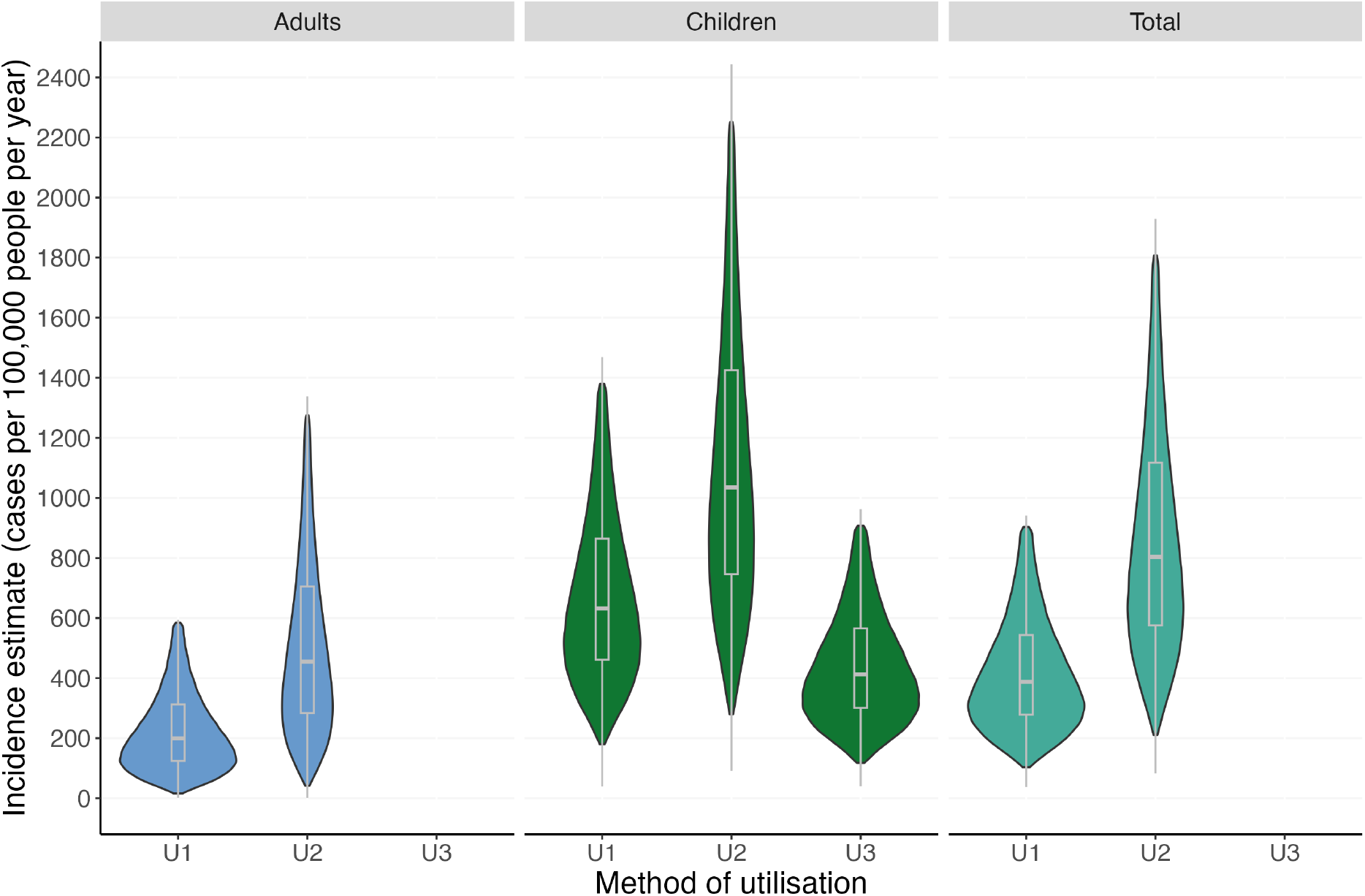
Posterior distributions of human brucellosis incidence estimates for the Ngorongoro Conservation area, 2016-2017, comparing estimates obtained using different estimates of healthcare utilisation. The 95% highest posterior density intervals and distributions for each estimate are presented in violins, with the boxplots indicating the median estimate and upper and lower quartiles. Estimates are presented for adults aged above 15 years (blue, left facet), children aged 15 years or below (green, middle facet), and the combined total population (teal, right facet). Within each facet (and population) estimates obtained using alternative healthcare utilisation parameters U1, U2 and U3 are compared.

## Discussion

We integrated healthcare centre based diagnostic testing results, test sensitivity and specificity estimates, and healthcare utilisation information in a Bayesian framework to provide estimates of brucellosis incidence in a northern Tanzanian pastoralist community. Our estimates indicate higher brucellosis incidence in the total population of the NCA compared to most but not all previous estimates from other global regions (15, 33). Importantly, our estimates account for low levels of healthcare utilisation and recognise the uncertainty of key parameters, leading to estimates that are likely to be more accurate and realistic than those generated through previous studies that have not accounted for these factors.

Given uncontrolled brucellosis in livestock, livestock management practices that lead to frequent human and livestock interaction, and cultural practices that increase animal to human *Brucella* spp. transmission risk, such as drinking unboiled milk (32), this population could plausibly experience a higher incidence of brucellosis than many others around the world. Livestock management and cultural differences are a likely explanation for the substantially higher incidence observed in this NCA population as compared to the nearby urban population of Moshi. An estimated incidence of 33 cases per 100,000 people per year was generated for the population around Moshi (18). Using methodology comparable to that applied for that previous study (our model simplification 5) (18), the equivalent value for the NCA population was 307.4 cases per 100,000 people per year, a nearly ten-fold difference. The incidence estimates obtained through our study highlight the need for improved surveillance for human brucellosis and argue that more widespread use of novel surveillance approaches that rely less on data generated at healthcare facility level is necessary, especially among high-risk communities such as pastoralists and others with low levels of healthcare utilisation.

Estimates of human brucellosis incidence vary widely by country and population, but they often range from 50 to 100 cases per 100,000 people per year in endemic areas (15). Notably, existing higher estimates range from 130 to as many as 583 cases per 100,000 people per year (15, 33). One recent estimate using serological data in northern Kenya estimated a seroincidence of approximately 6,000 infections (seroconversions) per 100,000 people per year (57), suggesting very frequent exposure and seroconversion in some communities, of which a proportion will result in clinical disease and presentation to healthcare facilities. Another study from a pastoralist community in Kenya estimated an incidence of 84 cases per 100,000 people per year in the total population, which was lower than the estimates presented here (16). Overall, there are a small number brucellosis incidence estimates from Africa, and substantial differences in the diagnostic test methods used to identify cases and the methods to derive incidence, greatly complicating the comparability of findings (15).

In addition to being higher than many existing estimates generated from different contexts and using highly variable methodologies, the brucellosis incidence values presented here are also highly uncertain, with wide density intervals. Rather than being a fault, the uncertainty of our estimates is likely realistic because it describes our state of knowledge about the true values of many key parameters and the level of confidence that these limited data can provide more clearly than simpler modelling methods. Some of the uncertainty in these models comes from the uncertain estimates for utilisation, which would likely have been more certain if the healthcare utilisation study had not been interrupted by the COVID-19 pandemic and the target sample size achieved. The explicit comparison between the full model and MS1 allows the scale of this effect to be assessed. Some of the uncertainty in our estimates also reflects the small number of brucellosis cases detected in the earlier prevalence study, a challenge that will always apply for low prevalence diseases. We would argue that our estimates reflect this uncertainty transparently and that superficially more precise or confident estimates generated from study designs and models that do not account for uncertainty in key parameters describe unrealistic systems and can be misleading in their presentation of apparent precision.

The brucellosis incidence estimates generated for children were much higher than those estimated for adults. This age structuring of disease is consistent with brucellosis incidence estimates in other parts of Tanzania that also found a higher incidence of brucellosis among children at some time points (18). In nomadic and semi-nomadic societies, children are reported to account for a high proportion of human brucellosis, and brucellosis is predominantly considered a paediatric health problem (11). Children may be at increased risk of acute brucellosis due to cultural practices such as drinking un-boiled milk and herding livestock (23, 58). Future studies could investigate how children specifically could be better protected from both infection and disease in this and similar endemic brucellosis contexts.

The results of the model simplification analysis clearly showed that accounting for low levels of healthcare utilisation markedly increased the incidence estimate obtained, and including uncertainty in the utilisation parameter (*U)* increased the HPDI. This is unsurprising, as not accounting for utilisation is effectively the same as assuming 100% of febrile persons access care, which is unlikely to be true. Estimates for healthcare utilisation for febrile illness in this study ranged from approximately 3% to approximately 20% depending on the population described (Table 1). A similar study in northern Tanzania found that, of 49 people presenting to the hospital with brucellosis-like symptoms, only 22% went to study health facilities within one month of symptom onset, with a median patient delay of 90 days (59). We recommend that any study aiming to accurately estimate disease incidence should carefully account for disease events missed due to lack of healthcare utilisation.

In this analysis, including test sensitivity and specificity parameters in the models had a lesser effect on the incidence estimates obtained. Accounting for sensitivity and specificity of the diagnostic tests produced only slightly different estimates from models assuming perfect test sensitivity and specificity, indicating that the two tests used in this study, SAT serology and blood culture, have high sensitivity and specificity when used in combination. Notably, the SAT and culture tests applied for the prevalence study were delivered through that research project and are not routinely available, and they have considerably improved sensitivity and specificity for detection of brucellosis cases compared to rapid tests for brucellosis that are more widely available (6).

Our comparison of different methods to estimate healthcare utilisation indicates that the design of healthcare utilisation surveys and their analyses also substantially impact the incidence estimates obtained through these methods. The lower values for actual rather than hypothetical care seeking behaviour (*U2* vs *U1*) suggests that estimates of this parameter based on indicative behaviour may lead to overestimates of *U* (and consequently under-estimates of incidence) when asking about hypothetical scenarios rather than actual past behaviour. The higher values of care seeking for children specifically in the scenario describing ‘brucellosis-like illness’ as compared to short duration febrile illness (*U3* vs *U1* and *U2*), and the resulting lower incidence estimates suggest that incidence estimates based on care seeking for short duration febrile illness only may lead to overestimates of brucellosis incidence. These results illustrate that the variation in the methods and exact phrasing of questions used to estimate healthcare utilisation can generate substantially different values and consequently different incidence estimates. The model simplification analysis shows that accounting for imperfect healthcare seeking is crucial, but how estimates of healthcare utilisation are quantified requires careful thought and consideration of approaches appropriate for any given context. The survey methods used to obtain our healthcare utilisation methods did not account for referral practices and may therefore have underestimated the proportion of individuals who would have reached Endulen hospital (even if their initial actions were to seek care elsewhere). Overall, given the varied manifestations of brucellosis and complexities of care seeking behaviour, the best methods to obtain accurate data on care seeking in this and other contexts should be evaluated.

The methods applied in this study assume a common value of healthcare utilisation within each age group modelled and do not therefore account for any heterogeneity in healthcare utilisation within populations. For example, the brucellosis-like illness scenario described in *U3* is specified for a female child, but we have not examined the influence of sex on care seeking in this study. Distance from a health facility also often has marked impact on the probability of care seeking (60). As people living in more remote areas are potentially at increased risk of brucellosis and less likely to seek care, this bias may lead to under-estimates of incidence in this study. It is also plausible that other causes of variation in healthcare utilisation are correlated with variation in risk for brucellosis and many other diseases, potentially leading to systematic biases in incidence estimates. Further work is needed to evaluate and overcome any biases introduced through these methodological choices.

An important caveat of our study is that it captured only brucellosis presentations that included febrile illness. Brucellosis cases not presenting with fever are not accounted for either in the prevalence study enrolment, which included only patients presenting with fever or having reported a fever in the past 72 hours, or the healthcare utilisation survey, which asked only about what would be or was done if a household member had a fever. These questions and approaches were designed to focus on acute presentations of brucellosis. However, since approximately 78% of people with clinical brucellosis present with fever (2), the prevalence study enrolment criteria likely excluded some individuals with clinical brucellosis presenting with symptoms other than fever, such as malaise, headache, or joint pain (61). The diagnostic approaches applied in the prevalence study were also focused on acute presentations and did not include investigations targeted to detect chronic disease manifestations. Chronic brucellosis can be debilitating and likely poses an additional public health burden not captured by the incidence estimates in our study, which is biased towards acute case detection. Therefore, although high, the incidence values presented here are plausibly under-rather than over-estimates for the total incidence of brucellosis in this population.

The human brucellosis incidence estimates presented for this pastoralist population in northern Tanzania are higher than those reported for other populations in Tanzania and indeed higher than most existing global estimates, revealing that brucellosis is an important health problem in this population. Our results show that accounting for low levels of healthcare utilisation is essential to avoid under-estimating disease incidence. These results are relevant to other contexts with extensive livestock management and endemic livestock brucellosis in East Africa, where high human brucellosis disease burdens and low levels of care seeking likely combine. These findings highlight the need for improved surveillance approaches. Novel methods for identifying and then treating brucellosis, such as efforts to reach people where they live rather than requiring them to travel to a distant health centre, are likely essential in populations such as this, where healthcare utilisation is low. Coordinated investment in increasing access to accurate brucellosis diagnostics and effective treatment, nationally in Tanzania and indeed regionally in East Africa, could lead to improved understanding of brucellosis epidemiology and better prevention and treatment of disease for many people living in at-risk communities.

## Data Availability

The minimal data sets and accompanying code are available at the Enlighten research data repository of the University of Glasgow via https://doi.org/10.5525/gla.researchdata.2317 (Healthcare utilisation study data) and https://researchdata.gla.ac.uk/978/ (Prevalence study data).

https://researchdata.gla.ac.uk/978/

## Author contributions

Holmes: Methodology, Formal analysis, Investigation, Writing - Original Draft, Writing - review and editing

De Glanville: Conceptualisation, Methodology, Formal analysis, Investigation, Writing - Original Draft, Writing - review and editing

Katiti: Investigation, Writing - review and editing

Mendes: Investigation, Writing - review and editing

Bodenham: Investigation, Writing - review and editing

Lukambagire: Investigation, Writing - review and editing

Mmbaga: Conceptualisation, Investigation, Funding acquisition, Writing - review and editing

Rubach: Conceptualisation, Investigation, Methodology, Funding acquisition, Writing - review and editing

Crump: Conceptualisation, Investigation, Methodology, Funding acquisition, Writing - review and editing

Cleaveland: Conceptualisation, Investigation, Methodology, Funding acquisition, Writing - review and editing

Viana: Methodology, Formal analysis, Writing - Original Draft, Writing - review and editing

Halliday: Conceptualisation, Methodology, Investigation, Writing - Original Draft, Funding acquisition, Writing - review and editing

## Acknowledgements

We thank the study teams and participants for both the prevalence and healthcare utilisation studies for their assistance in data collection. We also thank Tanzania Wildlife Research Institute (TAWIRI) and Ngorongoro Conservation Area Authority (NCAA) for approvals to conduct this project within the Ngorongoro Conservation Area.

